# sLOX-1 as a differential diagnostic biomarker for acute pulmonary embolism

**DOI:** 10.1101/2023.11.01.23297946

**Authors:** Jianing Wu, Wei Rong, Ke Ma, Jie Ma, Yuhong Mi, Zhi-Cheng Jing, Hui Zhang, Ping Li, Jie Du, Yulin Li

## Abstract

**Objective:** Diagnosing acute pulmonary embolism (PE) is challenging because of nonspecific clinical symptoms. Soluble lectin-type oxidized low-density lipoprotein receptor (sLOX-1) has differential expression in arterial and venous disease. This study aimed to evaluate the relevance of soluble lectin-type oxidized low-density lipoprotein receptor (sLOX-1) as a diagnostic biomarker for acute PE.

**Methods:** This observational study was performed at Beijing Anzhen Hospital in China. Patients with PE, aortic dissection (AD), myocardial infarction (MI) and healthy controls were enrolled in this cross-sectional study (n=90 each). Moreover, 730 patients with suspected PE were enrolled in this prospective study. The diagnostic performance of sLOX-1 was assessed using the receiver operating characteristic curve analysis.

**Results:** In the development set, sLOX-1 levels were significantly lower in patients with PE than in those with AD, MI, or healthy controls. In the validation cohort, the area under the curve (AUC) of sLOX-1 for patients with PE from other chest pain diseases, particularly from AD was significantly higher than that of D-dimer (ΔAUC=0.32; 95%CI, 0.26-0.37; P<0.0001) with 77.0% specificity and 74.5% positive predictive value at the threshold of 600 pg/mL derived from the development set. By integrating sLOX-1 into existing diagnosis strategy (Wells rules combined D-dimer), the number of patients who were further categorized as workup for PE decreased from 417 to 209, with the positive detection rate of computed tomographic pulmonary angiography increased from 35.1% to 67.0%. Six patients with PE were missed in 208 excluded patients at a failure rate of 2.88%.

**Conclusions:** Plasma sLOX-1 is a novel diagnostic tool that can rapidly categorize suspected PE as a workup for PE based on existing diagnostic strategy.

## Introduction

Pulmonary embolism (PE) is the most serious form of venous thromboembolism (^1^) and has high mortality rates (^2^). PE’s clinical symptoms are nonspecific, making the diagnosis difficult; therefore, advanced imaging with computed tomographic pulmonary angiography (CTPA) is required for a conclusive diagnosis. However, this technique cannot be performed for all patients with compatible symptoms owing to radiation and contrast exposure, as well as costs and equipment availability.

To address this limitation, guidelines have promoted algorithms that allow rapid, affordable, and large-scale diagnostic standardization. According to the 2019 European Society of Cardiology guidelines (^3^), the Wells rule was recommended to define if the pre-test probability of PE is unlikely (Wells score ≤ 4) or likely (Wells score≤4). For patients with a low likelihood of PE, the decision to perform CTPA was determined by additional D-dimer level evaluations, whilst patients with an unlikely probability of a positive D-dimer test result underwent CTPA (^3^). Even so, <30% patients were finally diagnosed with PE in all patients undergoing CTPA (^4^). Therefore, further workup is necessary to better evaluate the likelihood of PE in patients.

LOX-1 (lectin-type oxidized low-density lipoprotein [oxLDL] receptor) has emerged as a promising target for the prediction of cardiovascular risk. Accumulating evidence supports its involvement not only in oxLDL internalization but also in endothelial dysfunction, atherosclerosis, plaque instability, and thrombogenesis (^5^). Elevated soluble LOX-1 (sLOX-1) concentrations have been observed in coronary artery disease (CAD) (^6^), myocardial infarction (MI) (^6,7^), and aortic dissection (AD) (^8^). PE is an embolic disease occurring in the veins, which is different from coronary artery embolism in CAD and aortic intramural thrombosis in AD. Furthermore, our reanalysis of the GEO database GSE43475 (^9^) revealed that the mRNA expression of LOX-1 in the femoral and iliac veins was significantly lower than that in the femoral and iliac arteries. Additionally, sLOX-1 is reportedly generated by ectodomain shedding by (disintegrin and metalloproteinase with thrombospondin motifs) ADAMTS enzymes (^10,11^). ADAMTS13 activity decreased in patients with venous thromboembolism (^12^), whereas the activity and level of ADAMTs increased in patients with CAD and AD (^13–15^). Therefore, we hypothesized that sLOX-1 would be lower in PE, in contrast to being higher in AD and CAD, making it a promising marker for deciphering veinous thromboembolism and facilitating the categorization of patients with PE.

We aimed to determine whether plasma sLOX-1 can distinguish PE from other chest pain diseases and can help rapidly categorize patients requiring PE workup based on existing diagnostic strategies.

## Materials and Methods

### Study design and setting

This observational study was performed at Beijing Anzhen Hospital in China. Two separate populations were enrolled: the development set, for assessing whether sLOX-1 differed between PE and other chest pain groups and HC in a cross-sectional study, and the validation cohort, for evaluating the diagnostic value of sLOX-1 in a prospective study. The study was conducted according to the Strengthening the Reporting of Observational Studies in Epidemiology equator checklist (^16^).

### Selection of Participants

Participants were consecutive patients admitted to Beijing Anzhen Hospital of Capital Medical University (Beijing, China). The development set was enrolled between January 2016 and January 2017, where the patients with confirmed PE clinically were age-and sex-matched with the confirmed AD, MI, and healthy control (HC) groups (n=90 per group). Detailed disease diagnosis definitions are available in the *Supplementary Material*.

For the validation cohort, consecutive patients aged >18 years who presented to the emergency department from January 2017 to December 2018 were eligible if they had clinical suspicion of PE (first or recurrent), defined as an acute onset or worsening shortness of breath or chest pain without any obvious etiology. Patients were included only if PE was considered as a differential diagnosis by the attending physician. Patients were excluded if they were receiving anticoagulant therapy for another indication (e.g., atrial fibrillation), or if they had an allergy to the contrast medium, impaired renal function, life expectancy <3 months, or ongoing pregnancy. Clinical decisions were made by the attending physicians irrespective of study participation.

Information on demographic characteristics, history, clinical presentation, physical examination, imaging, and management was obtained from medical records. All patients underwent standard surgical or medical treatments and provided written informed consent. This study was approved by the Institutional Review Board of Beijing Anzhen Hospital (No. 2018048X). The study design was conducted in accordance with the Declaration of Helsinki. (ClinicalTrials, NCT04118634).

### Case definition and adjudication

PE diagnosis was made by two expert physicians and was defined as the evidence of PE on CTPA. The study was dichotomous in nature, considering that PE was considered absent on the basis of negative CTPA results. If such data were unavailable, the adjudication was considered clinical. PE was considered absent if: (i) an alternative diagnosis (AltD) was available and (ii) the participants had an uncomplicated clinical course, or received an AltD during the follow-up period. The adjudicators conferred and reconsidered their classification, and requested consultation from a third-blinded adjudicator.

### Follow-up

Patients without conclusive diagnostic data were registered for clinical follow-up for case adjudication. After 14 days, the patients or family members were interviewed via telephone using a structured questionnaire. The following events were queried: diagnosis of PE, AltD, or death.

### Sample size calculation

The sample size of the development set was based on the assumption that sLOX-1 had a sensitivity of 90% and a specificity of 60%. Using power (1-beta) = 95% and alpha = 0.05, the sample sizes for sensitivity and specificity were 53 positive participants and 82 negative participants, respectively. The sample size was calculated using the PASS 15 Power Analysis and Sample Size Software (2017). NCSS, LLC. Kaysville, Utah, USA.

For the validation cohort, we included 730 patients to provide accurate estimates, focusing on the exclusion of PEs with a minimum number of missed cases. We assumed failure rate of the integration diagnostic rule-out strategy (Wells score≤4/D-dimer-/sLOX-1-) at 0.8%, which is based on reported failure rate of existing diagnostic tools for PE (^17^). Our study aimed to test the null hypothesis that the failure rate of the indicated diagnostic tools exceeds 3%. Using a one-sided Type I error of 0.05 and a Type II error of 0.2, we needed to exclude approximately 262 participants to reject the null hypothesis.

### Statistical analysis

General characteristics were assessed using means and standard deviations, whilst medians and interquartile ranges were used for continuous variables, and proportions were used for categorical variables. Differences between any two groups were compared using a two-sample t-test (for normally distributed continuous variables), the Mann–Whitney U test (for non-normally distributed continuous variables), or Fisher’s exact or chi-squared tests (for categorical data). Significant differences in plasma sLOX-1 and D-dimer levels across the different diagnostic groups were adjusted for multiple comparisons using Dunnett’s correction. Multivariate logistic regression was used to analyze the independent predictors of PE.

To evaluate diagnostic performance, sensitivity, specificity, negative/positive predictive values, and likelihood ratios, receiver operating characteristic (ROC) curves were obtained. The optimal cutoff value for plasma sLOX-1 levels was established using the Youden index. Significant differences between AUCs were examined using DeLong statistics. A Fagan nomogram was developed to visualize the effect of sLOX-1 findings on the probability of PE.

All statistical analyses were performed using Graphad Prism (GraphPad Software, San Diego, CA, USA) and R version 4.0.3 (R Foundation for Statistical Computing, Vienna, Austria), specifically, the “pROC” (version 1.18.0) and “tableone” (version 0.13.2) packages. All p-values were two-tailed, and statistical significance was defined as P<0.05.

## Results

### 1. Plasma sLOX-1 and D-dimer comparisons among different diagnostic groups within the development set

The baseline demographic and clinical characteristics of the 360 patients in the development set are shown in ***Supplementary Material: Table S1***. Plasma sLOX-1 and D-dimer levels in the diagnostic and HC groups are shown in ***Supplementary Material: Figure S1***. The level of plasma sLOX-1 was significantly lower in PE (median, 168.90 pg/mL) compared to AD (median, 1185.56 pg/mL; adjusted P<0.0001), MI (median, 891.47 pg/mL; adjusted P<0.0001) and HC (median, 475.13 pg/mL; adjusted P<0.0001) groups. Plasma D-dimer concentrations were elevated in patients with AD, MI, and PE compared with HC. There was no significant difference in the D-dimer levels between the AD and PE groups. These results suggest that sLOX-1 levels may differ between venous and arterial thromboembolisms.

### 2. AUCs of plasma sLOX-1 and D-dimer for PE vs. other chest pain diseases and HC within the development set

The AUCs for PE vs. other diagnostic groups are shown in ***Supplementary Material: Figure S2*,** and the corresponding sensitivities and specificities are reported in ***Supplementary Material: Table S2***. The overall AUC of sLOX-1 for PE vs. MI and AD was significantly higher than that of D-dimer (ΔAUC, 0.11 [95% CI, 0.04–0.17]; P=0.001), with an optimal sLOX-1 threshold of 594.6 pg/mL. Moreover, the AUC of the sLOX-1 was superior to that of the D-dimer when only AD patients were compared with PE patients (ΔAUC, 0.22 [95% CI, 0.12–0.32]; P<0.0001), with the optimal sLOX-1 threshold of 594.9 pg/mL. The AUC of plasma sLOX-1 to distinguish PE from MI was similar to that of D-dimer (ΔAUC, 0.002 [95% CI, -0.06 – 0.06]; P=0.95). The AUC of plasma sLOX-1 to distinguish PE from HC was inferior to that of D-dimer (ΔAUC, -0.22 [95% CI, -0.28 – -0.15]; P<0.0001). Based on these findings, sLOX-1 at around 600 pg/mL was the optimal threshold for distinguishing PE from the other chest pain diagnoses (MI plus AD).

### 3. Plasma sLOX-1 and D-dimer comparisons among different clinical diagnostic groups within the validation cohort

Seven hundred and thirty patients with suspected PE were included in the validation cohort (**Figure 1A**). Diagnostic workup details are presented in ***Supplementary Material: Figure S3***. PE was adjudicated in 146 patients. In remaining patients, the following alternative diagnoses were considered: AD (209 patients, 28.60%), MI (290, 39.70%), and others (85, 11.60%). **Table 1** shows the clinical characteristics of the 146 PE patients and 584 non-PE patients. Plasma sLOX-1 and D-dimer level among diagnostic groups are shown in **Figure 1B, C**. Median sLOX-1 levels in patients with PE were 12.1-fold, 8.0-fold, and 7.7-fold lower than in patients with AD, MI, and other diagnoses, respectively. D-dimer levels in patients with PE were significantly higher than in patients with MI and other diseases, but not in patients with AD.

**Figure 1.**
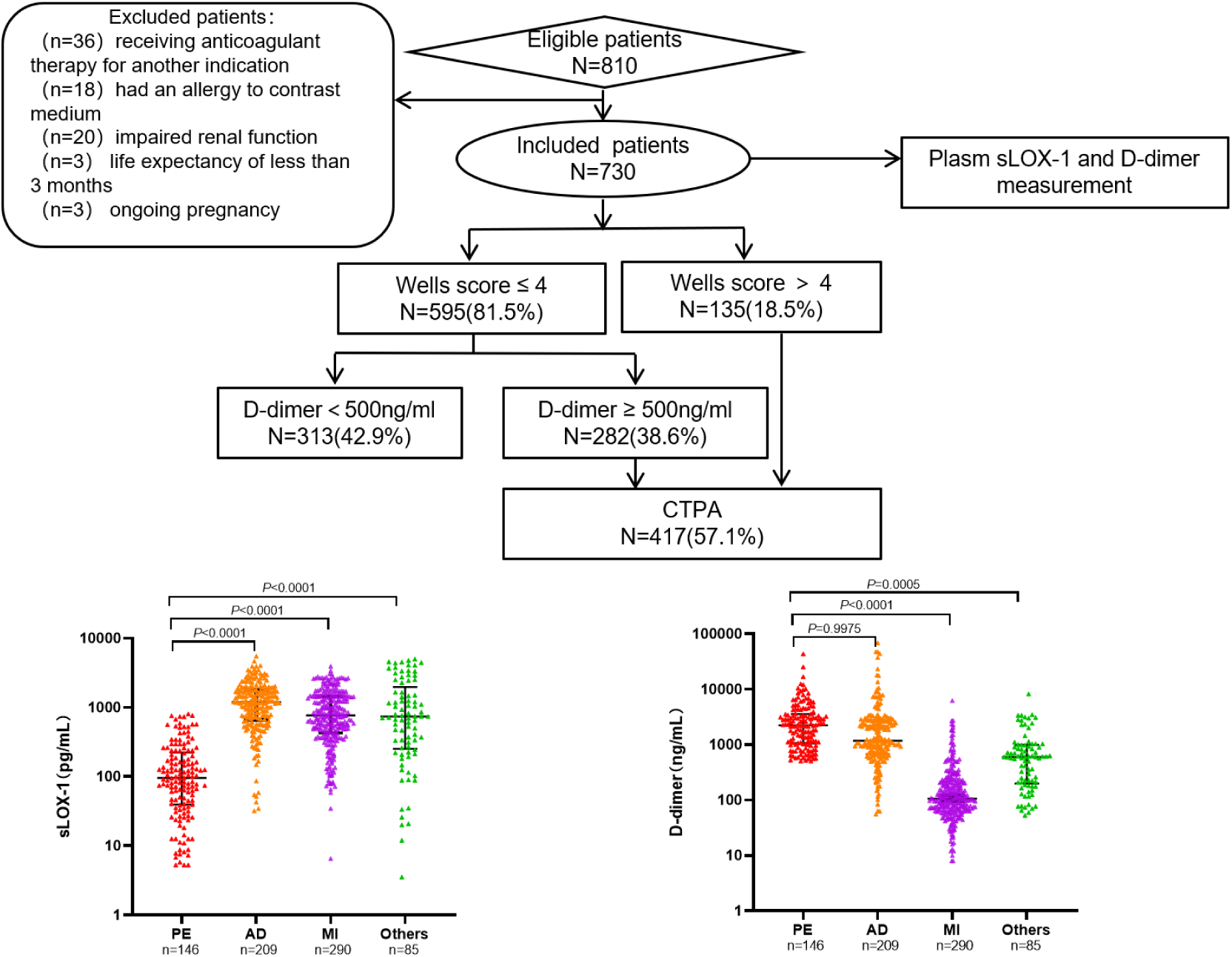
Study flow diagram and levels of plasma biomakers in the validation cohort. (A) A flow diagram of the study in the validation. Study population and the results of Wells score and D-dimer test in patients with suspected PE. % refers to 730 study patients. (B) Plasma sLOX-1 and D-dimer comparisons among different clinical diagnostic groups in the validation cohort. Biomarker concentrations are shown as median ± interquartile range. P-values are Dunnett corrected for multiple comparisons. Abbreviations: PE, pulmonary embolism; AD, aortic dissection; MI, myocardial infarction; Others, other chest pain diseases with no definite diagnosis; CTPA, computed tomographic pulmonary angiography.

**Table 1.**
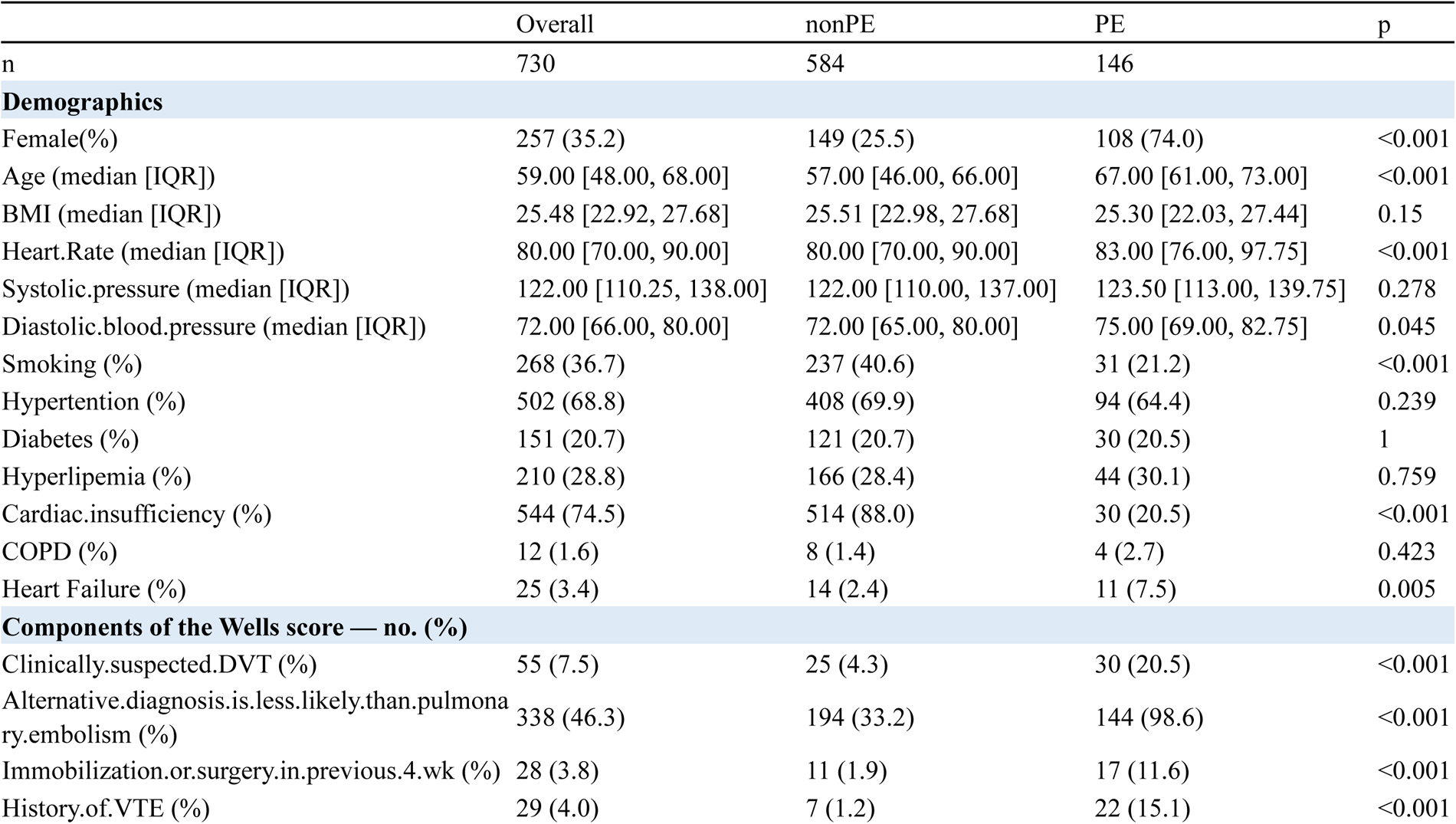
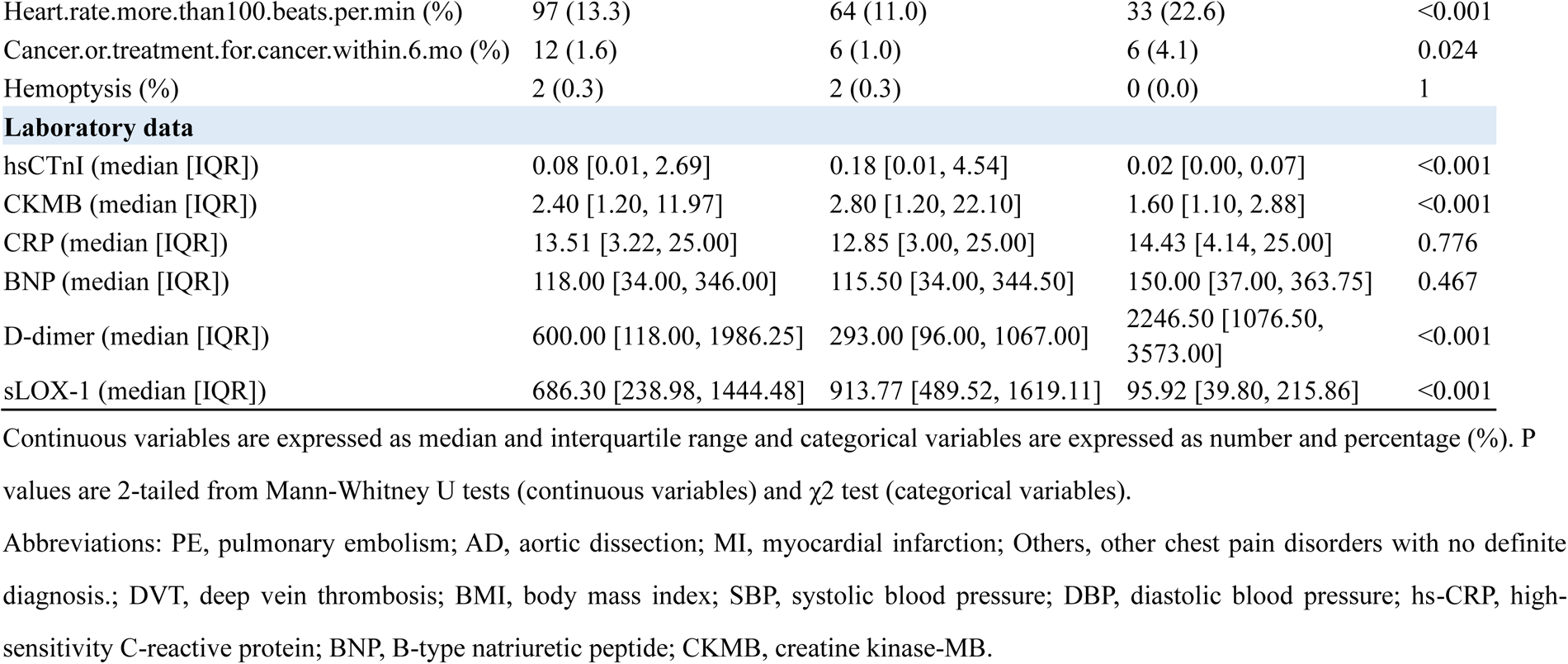
Baseline characteristics of validation cohort.

### 4. Diagnostic performance of plasma sLOX-1 within the validation cohort

Among 730 patients with suspected PE, sLOX-1 had a significantly higher overall AUC for PE vs. non-PE (AD, MI, and other diagnoses) than D-dimer (**Figure 2**). At the thresholds of 600 pg/mL for sLOX-1 and 500 ng/mL for D-dimer, sLOX-1 had higher specificity, positive predictive value (PPV), and positive likelihood ratio (LR+) than D-dimer (**Table 2**). Analysis of sLOX-1 and D-dimer diagnostic performance in different subgroups showed that sLOX-1 had higher AUC, specificity, PPV, and LR+ than D-dimer in patients with PE vs. AD and PE vs. AD plus MI. Specifically, the specificity of sLOX-1 was significantly higher than that of D-dimer (77.0% vs. 17.2%, respectively) in patients with PE vs. AD. The PPV (74.5%) and LR^+^ (4.2) were also significantly higher than those of D-dimer (45.8% and 1.2, respectively). However, the performance of sLOX-1 was inferior to that of D-dimer in distinguishing PE from MI alone.

**Figure 2.**
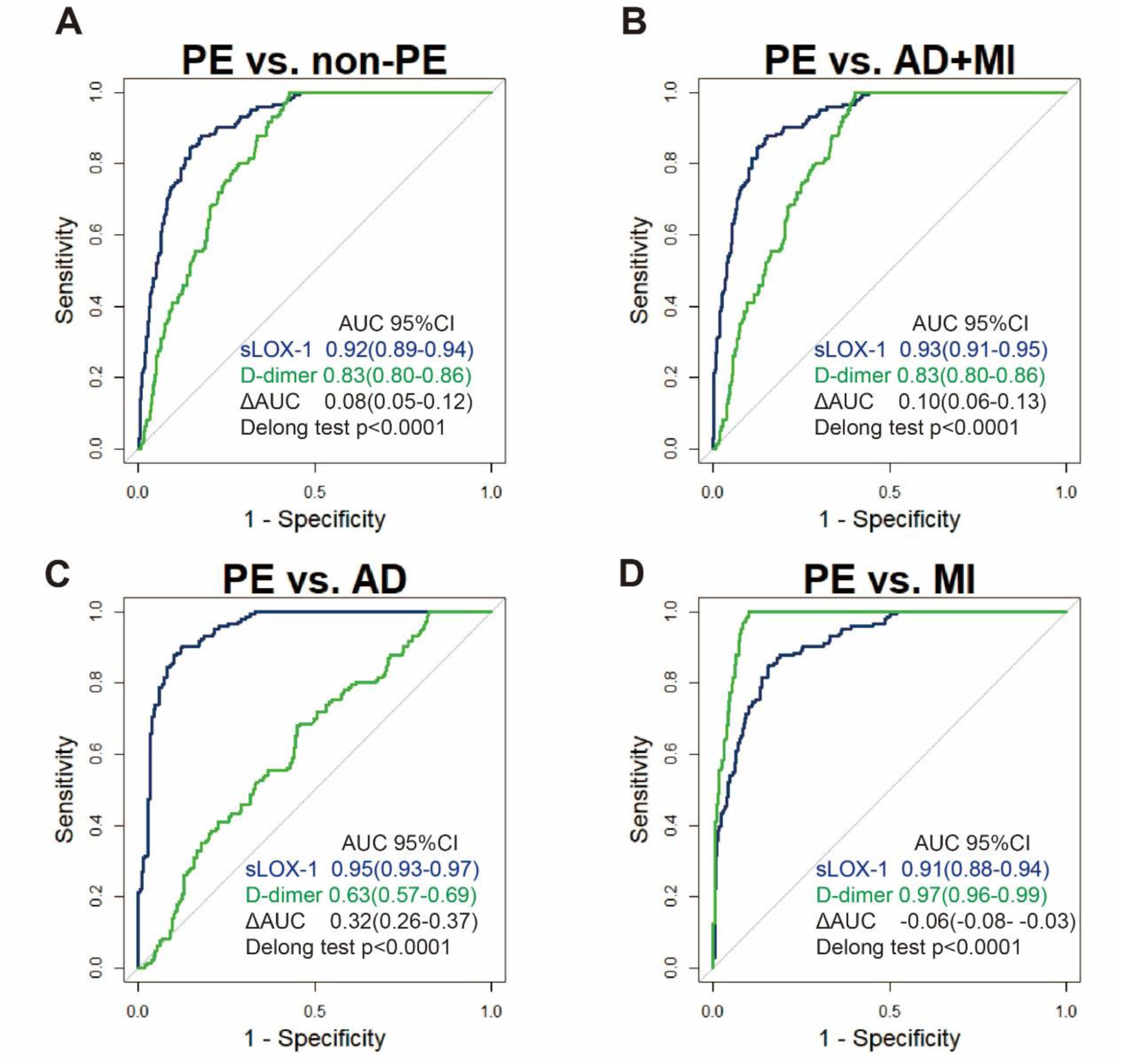
ROC curve analyses of plasma sLOX-1 and D-dimer for PE vs. other chest pain diseases in the validation cohort. Receiver-operating-characteristic area under the curve (AUC) of sLOX-1 and D-dimer levels in differentiating PE from non-PE (A), AD and MI (B), AD (C), and MI (D). The gray diagonal lines represent the results expected by chance. Abbreviations: PE, pulmonary embolism; AD, aortic dissection; MI, myocardial infarction; non-PE, AD and MI and other chest pain diseases with no definite diagnosis.

**Table 2.** Diagnostic performances of plasma sLOX-1 and D-dimer for differentiating PE from other chest pain diseases in the validation cohort.

|  | Sensitivity | Specificity | NPV | PPV | LR+ | LR- | Cut-off |
| --- | --- | --- | --- | --- | --- | --- | --- |
| **PE vs. non-PE** |  |  |  |  |  |  |  |
| sLOX-1 | 0.959 | 0.663 | 0.985 | 0.415 | 2.843 | 0.062 | 600pg/ml |
| D-dimer | 1.000 | 0.572 | 1.000 | 0.369 | 2.336 | 0.000 | 500ng/ml |
| **PE vs. AD+MI** |  |  |  |  |  |  |  |
| sLOX-1 | 0.959 | 0.675 | 0.983 | 0.464 | 2.954 | 0.061 | 600pg/ml |
| D-dimer | 1.000 | 0.597 | 1.000 | 0.421 | 2.483 | 0.000 | 500ng/ml |
| **PE vs. AD** |  |  |  |  |  |  |  |
| sLOX-1 | 0.959 | **0.770** | 0.964 | **0.745** | **4.175** | 0.053 | 600pg/ml |
| D-dimer | 1.000 | 0.172 | 1.000 | 0.458 | 1.208 | 0.000 | 500ng/ml |
| **PE vs. MI** |  |  |  |  |  |  |  |
| sLOX-1 | 0.959 | 0.607 | 0.967 | 0.551 | 2.439 | 0.068 | 600pg/ml |
| D-dimer | 1.000 | 0.900 | 1.000 | 0.834 | 10.000 | 0.000 | 500ng/ml |

Sensitivity, specificity, PPV, NPV, LR+, LR- were calculated using the best cut-offs of 600pg/mL for sLOX-1 and 500ng/mL for D-dimer. LR+, positive likelihood ratio; LR-, negative likelihood ratio; PPV, positive predictive value. NPV, negative predictive value.

The Wells score and D-dimer levels were used to evaluate clinical probability for PE diagnosis. Of the 730 patients, 313 patients with a Wells score ≤ 4 (“unlikely” clinical probability) and negative D-dimer value (D-dimer < 500 ng/mL) were considered non-PE and excluded from further testing, while remaining 417 patients were recommended CTPA (**Figure 1A**). For patients recommended CTPA, sLOX-1 specificity was 75.3% in 282 patients, and 71.2% in the group with 135 patients. In addition, the PPV of sLOX-1 was 52.2% in the group with 282 patients, and 84.4% in the group with 135 patients. However, for the group with a negative D-dimer value requiring no further testing, sLOX-1 specificity was only 40.9% **(*Supplementary Material: Table S3*)**.

### 5. Additive value of sLOX-1 based on existing diagnostic strategy

In multivariable logistic regression analysis, sLOX-1 was an independent predictor of PE along with female sex, age and other risk factors related to PE. (***Supplementary Material: Table S4***). Moreover, ROC analysis showed that the integration of sLOX-1 with the Wells score and the D-dimer in 282 patients and integration of sLOX-1 with Wells score in 135 patients significantly increased the diagnostic accuracy for PE (**Figure 3A and C**). Among the 282 patients with an “unlikely” clinical probability with a positive D-dimer value, the prior probability of PE was 22.3% (63/282); while, the detection of positive sLOX-1 led to a posterior probability of 52.2% (59/113).

**Figure 3.**
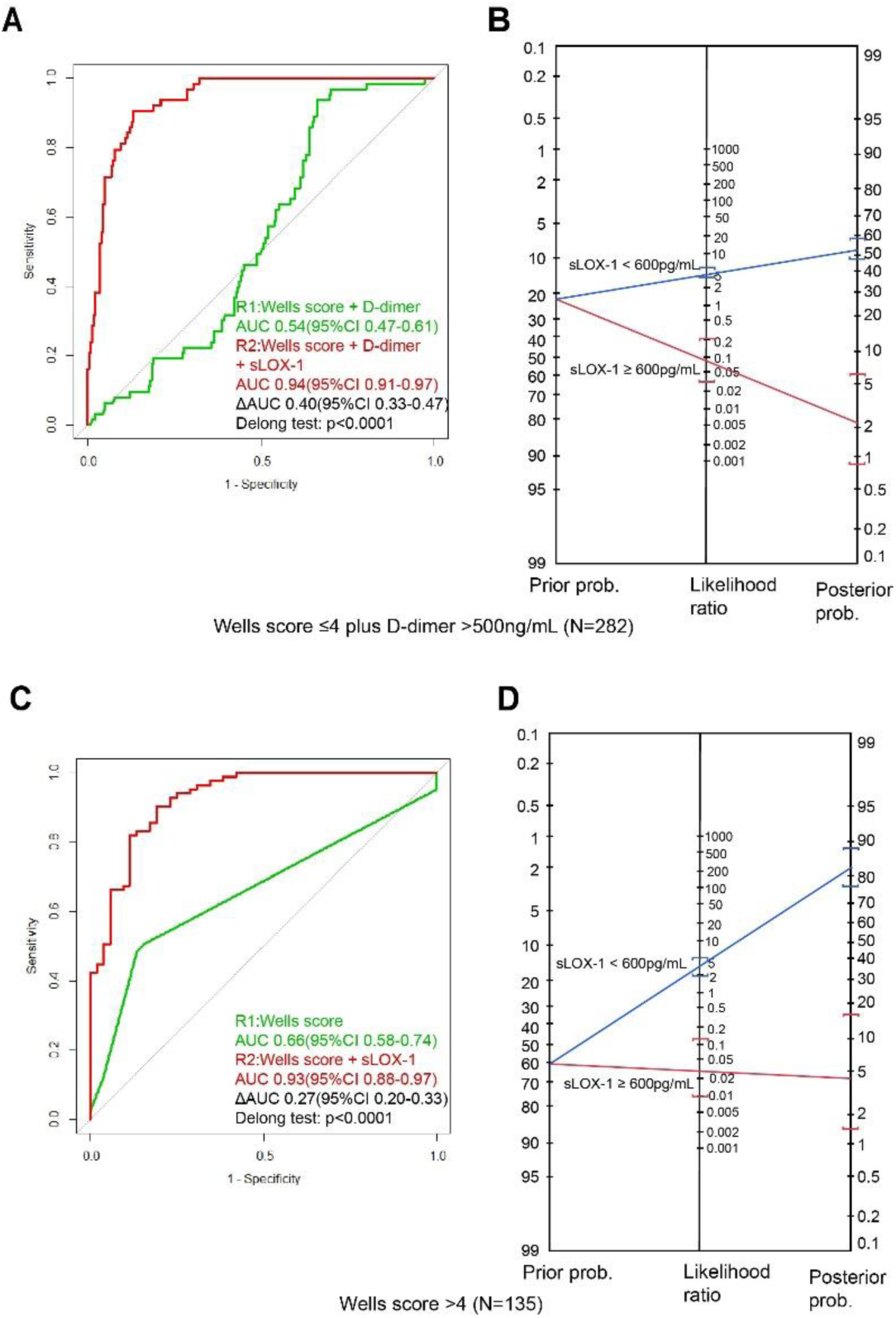
Additive diagnostic value of sLOX-1 to Wells score with D-dimer. (**A**) ROC curves for diagnosis of PE of the Wells score with D-dimer (green line), Wells score with D-dimer plus sLOX-1(red line). **(B)** Fagan nomogram showing the additive effect of sLOX-1 to Wells score with D-dimer in 282 patients with unlikely probability and positive D-dimer. **(C)** ROC for diagnosis of PE of the Wells score (green line) and Wells score plus sLOX-1 (red line). **(D)** Fagan nomogram showing the additive effect of sLOX-1 to Wells score in 135 patients with likely probability. The clinical probability of PE is displayed on the left as “Prior Prob.” The middle line represents the result of sLOX-1. When a straight line is drawn through the prior probability and sLOX-1 result, the post-test probability of PE is found on the right line (“Post Prob”).

Contrarily, the absence of a positive sLOX-1 result led to a posterior probability of 2.37% (4/169) (**Figure 3B**). Among the 135 patients with a “likely” clinical probability, the prior probability of PE in this group was 61.5% (83/135); however, the detection of positive sLOX-1 led to a posterior probability of PE of 84.4% (81/96). Contrarily, the absence of a positive sLOX-1 results led to a posterior probability of 5.13% (2/39) (**Figure 3D**).

### 6. sLOX-1 contributed in rapid categorization of patients for PE workup on the basis of existing diagnostic strategy

In the existing diagnostic strategy, 417 patients were categorized as requiring a workup for PE, which eventuated in 146 being diagnosed as positive by CTPA. By adding sLOX-1 to the existing diagnostic strategy, 209 patients were more efficiently categorized as requiring workup for PE. The results found that 140 PEs were detected positive, meaning that the positive detection rate of CTPA increased from 35.1% to 67.0% (**Figure 4**). In the 208 patients that were additionally excluded from the study with a negative sLOX-1 value (≥600 pg/mL), six PE patients were missed with a failure rate of 2.88% (6/208) (***Supplementary Material: Table S5***).

**Figure 4.**
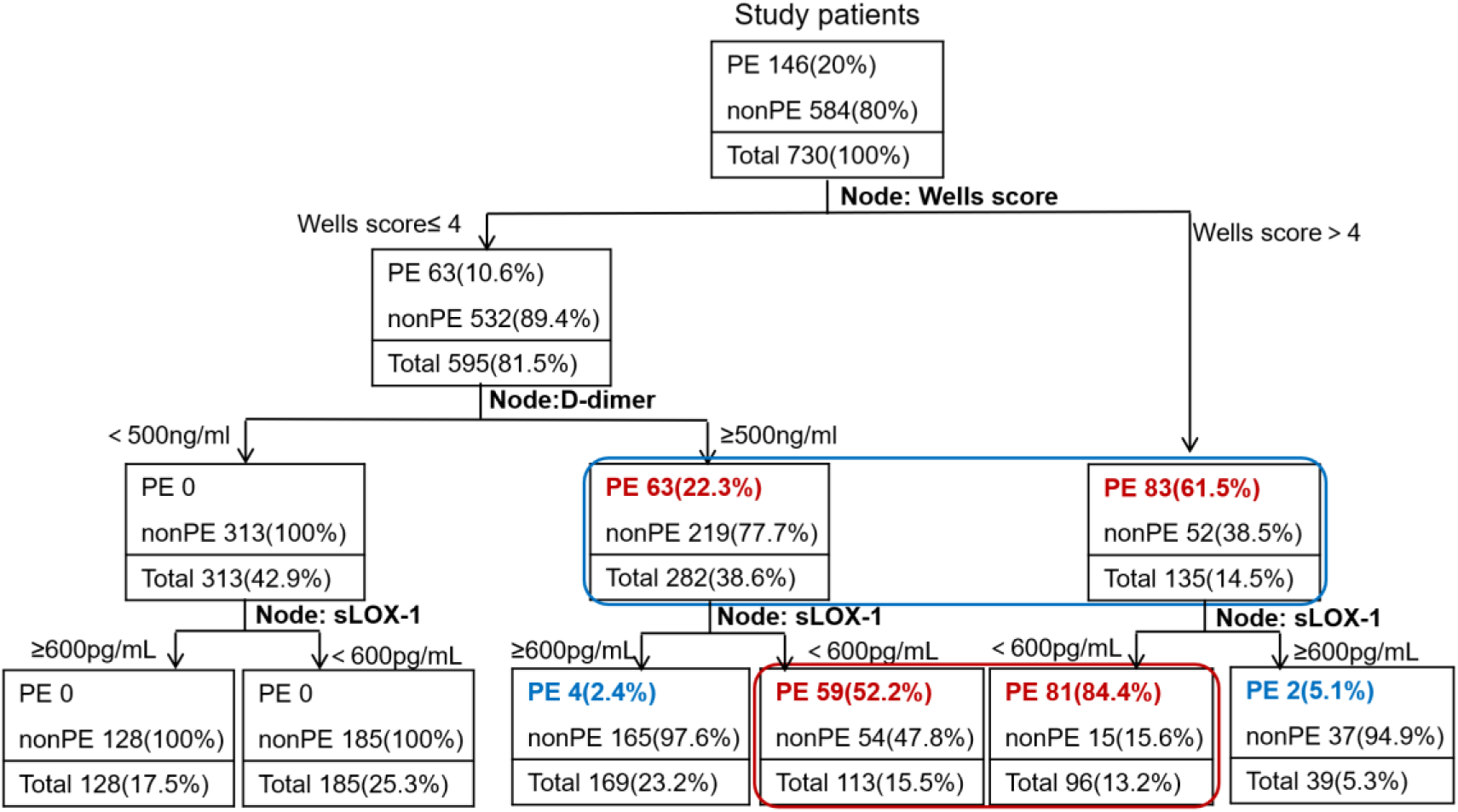
Decision-tree analysis. A total of 417 patients were categorized as requiring workup for PE based on the existing diagnostic strategy (Wells score>4 combined with D-dimer≥500ng/mL; blue box). A total of 209 patients were further categorized as requiring a workup for PE when sLOX-1 was added to the existing diagnostic strategy (red box). At the level of each node, the number of patients with PE or non-PE (including % within node) and the total number of patients (with % of study cohort) are shown.

## Discussion

This study analyzed data from 1090 participants, including 360 in the development set and 730 with suspected PE in the validation cohort. Results suggest that sLOX-1 is a potential diagnostic biomarker for distinguishing PE from other chest pain diseases, especially from AD. Adding sLOX-1 to existing diagnostic strategy further helped categorize patients requiring workup for PE.

Baseline characteristics of our study population were comparable to those reported in other population-based studies. For example, the average age (59 vs. 53 years), heartbeat (80 vs. 87 bpm), and D-dimer level (600 ng/mL vs. 700 ng/mL) of patients with suspected PE in our validation cohort were comparable to those in two previous studies (^18, 19^). The overall prevalence of PE in our study (20%) was also comparable to that reported in a previous multicenter study (19%) (^18^). This study provided the first evidence that sLOX-1 can specifically distinguish between PE and non-PE.

Our study has some advantages. First, this was a rationale-derived proof-of-concept study. Another strength of this study is the utilization of a cross-sectional design followed by a separate prospective cohort for validation. Initially, a cross-sectional design was implemented to test the hypothesis and establish the cutoff value for sLOX-1. Results indicated that plasma levels of sLOX-1 were elevated in both AD and MI patients when compared to HCs, which aligns with previous findings (^8, 20^). Notably, the level of sLOX-1 was lower in patients with PE than that in non-PE patients, and even below the levels observed in HC. This may be due to ADAMTS stimulating sLOX-1 cleavage at the cell membrane. The levels and activity of ADAMTS are reportedly diminished in venous thrombosis, which may explain the marked reduction in sLOX-1 in PE patients. For instance, ADAMTS-13 activity in patients with portal vein thrombosis (PVT) was significantly lower than in those without PVT (median 16.8 vs. 69.1 %) (^21^). Additionally, a case-control study found an excess of rare-coding single nucleotide variants in the ADAMTS-13 gene in patients with deep vein thrombosis, and such patients demonstrated lower plasma levels of ADAMTS-13 activity than patients without them (^22^).

The significant decrease in plasma sLOX-1 levels suggests that this protein may serve as a valuable differential diagnostic biomarker. To further explore this potential role, we conducted a prospective study to assess the differential diagnostic performance of sLOX-1 and its added value to existing diagnostic strategies for patients with suspected PE. While D-dimer is recognized as a diagnostic marker for PE due to its high sensitivity, it lacks specificity and cannot distinguish between acute AD and PE (^23^).

In our study, D-dimer exhibited superior sensitivity with 100% compared to sLOX-1 with 95.9%. However, sLOX-1 was more specific than D-dimer in differentiating between PE and non-PE cases (66% vs. 57%). We also found that copeptin, a diagnostic and prognostic (^24^) marker for PE, exhibited higher specificity (83.7%) (^25^) compared to sLOX-1 (66.3%) in our validation cohort. However, previous study had a relatively small population (n=90) and lacked a validation cohort and subgroup analysis (^25^). Notably, sLOX-1 demonstrated a superior performance to D-dimer in distinguishing between PE and AD patients, with a specificity of 77% compared to 17%.

Our decision-tree analysis suggests that incorporating sLOX-1 into the diagnostic workup for patients with suspected PE may improve the detection rate. The addition of sLOX-1 increased the positive detection rate of CTPA from 35% to 67%, compared with existing diagnostic strategy. However, six PE cases were missed, resulting in a failure rate of 2.88%, which was acceptable given that the failure rate of a PE diagnostic strategy should not exceed 3% (^26^).

Our study had some limitations. Firstly, as it was conducted in a single-center, this study was susceptible to patient selection bias. Therefore, the generalizability of our findings should be verified in a large-scale, prospective multicenter study. Secondly, the AUC of sLOX-1 was inferior to that of D-dimer in distinguishing PE from MI alone and from HC. This implied that sLOX-1 was mainly expected to distinguish PE from AD in the emergency room. Next, the detection of sLOX-1 relied on ELISA, which may not be suitable for large-scale applications. The absolute values of sLOX-1 concentrations may differ depending on the assay method used, which could affect the proposed diagnostic thresholds. Additionally, larger population studies are necessary to confirm our findings and assess the potential utility of sLOX-1 in clinical care.

In summary, our findings suggest that plasma sLOX-1 levels can improve diagnostic specificity for PE, address the limitations of D-dimer in both PE and AD patients, and enhance the diagnostic workup of patients suspected of having PE.

## Data Availability

All data referred to in the manuscript is availability.

## Author contributions

Yulin Li contributed to the study design. Jianing Wu and Wei Rong contributed to collection of blood sample and clinical information. Jianing Wu and Ke Ma performed data analyses. Hui Zhang contributed to statistical guidance. Jianing Wu wrote the manuscript. Yuhong Mi, Zhi-Cheng Jing and Ping Li contributed to clinical assessment. Yulin Li and Jie Du guided the study and approved the final version.

